# Anxiety and depression symptoms after COVID-19 infection: results from the COVID Symptom Study app

**DOI:** 10.1101/2021.07.07.21260137

**Authors:** Kerstin Klaser, Ellen J. Thompson, Long H. Nguyen, Carole H. Sudre, Michela Antonelli, Benjamin Murray, Liane S. Canas, Erika Molteni, Mark S. Graham, Eric Kerfoot, Liyuan Chen, Jie Deng, Anna May, Christina Hu, Andy Guest, Somesh Selvachandran, David A Drew, Marc Modat, Andrew T. Chan, Jonathan Wolf, Tim D. Spector, Alexander Hammers, Emma L. Duncan, Sebastien Ourselin, Claire J. Steves

**Affiliations:** School of Biomedical Engineering & Imaging Sciences, King’s College London, London SE1 7EU, UK; Department of Twin Research and Genetic Epidemiology, King’s College London, London SE1 7EH, UK; Clinical & Translational Epidemiology Unit, Massachusetts General Hospital and Harvard Medical School, 100 Cambridge Street, Boston, MA, USA; Division of Gastroenterology, Department of Medicine, Massachusetts General Hospital and Harvard Medical School, 100 Cambridge Street, Boston, MA, USA; MRC Unit for Lifelong Health and Ageing, Department of Population Science and Experimental Medicine, University College London, UK; Centre for Medical Image Computing, Department of Computer Science, University College London, UK; King’s College London & Guy’s and St Thomas’ PET Centre, London, UK; Zoe Limited, London SE1 7RW, UK

**Keywords:** COVID-19, SARS-CoV-2, Mental health, Anxiety, Depression, GAD2, PHQ2

## Abstract

**Background:** Mental health issues have been reported after SARS-CoV-2 infection. However, comparison to prevalence in uninfected individuals and contribution from common risk factors (e.g., obesity, comorbidities) have not been examined. We identified how COVID-19 relates to mental health in the large community-based COVID Symptom Study.

**Methods:** We assessed anxiety and depression symptoms using two validated questionnaires in 413,148 individuals between February and April 2021; 26,998 had tested positive for SARS-CoV-2. We adjusted for physical and mental pre-pandemic comorbidities, BMI, age, and sex.

**Findings:** Overall, 26.4% of participants met screening criteria for general anxiety and depression. Anxiety and depression were slightly more prevalent in previously SARS-CoV-2 positive (30.4%) vs. negative (26.1%) individuals. This association was small compared to the effect of an unhealthy BMI and the presence of other comorbidities, and not evident in younger participants (≤40 years). Findings were robust to multiple sensitivity analyses. Association between SARS-CoV-2 infection and anxiety and depression was stronger in individuals with recent (<30 days) vs. more distant (>120 days) infection, suggesting a short-term effect.

**Interpretation:** A small association was identified between SARS-CoV-2 infection and anxiety and depression symptoms. The proportion meeting criteria for self-reported anxiety and depression disorders is only slightly higher than pre-pandemic.

**Funding:** Zoe Limited, National Institute for Health Research, Chronic Disease Research Foundation, National Institutes of Health, Medical Research Council UK

## Introduction

Studies from previous coronaviruses suggesting an increased risk of neurological disorders^1^, and case studies^2-4^ and findings^5-7^ regarding the impact of SARS-CoV-2 infection on the central nervous system^8-11^ led to the hypothesis that anxiety/depression symptoms may be more prevalent in individuals after SARS-CoV-2 infection. Indeed, several reports suggest that COVID-19 survivors are at increased risk of mood and anxiety disorders three months post-infection^12-15^. Moreover, the Office for National Statistics reported a steep increase in anxiety/depression symptoms in the general public (irrespective of infection status) compared to pre-pandemic data, adjusting for socioeconomic factors^16^.

Quantifying the relationship of SARS-CoV-2 infection on anxiety/depression symptoms *per se* requires disentangling the consequences of infection from other factors such as lockdown measures. Direct links to health records may enable assessment of SARS-CoV-2 infection on psychiatric diagnoses^15^; however, it takes time and resources to acquire large cohorts for such longitudinal studies. Alternatively, analysis of self-reported real-time data allows for faster and timely insights into effects on mental health from SARS-CoV-2 infections.

This study aimed to assess prevalence of anxiety/depression symptoms in individuals with and without prior SARS-CoV-2 infection using a large community cohort, including assessment of other known mental health predictors. We used data from 413,148 tested non-healthcare workers who answered a mental health survey between February and April 2021 via the COVID Symptom Study app^17^.

## Methods

### Sample

Data were acquired by the COVID Symptom Study app^17^, a mobile application developed by health data company Zoe Limited in collaboration with King’s College London (KCL), the Massachusetts General Hospital, Lund University, and Uppsala University. The app was launched on 24 March 2020 and allows users to report their health status (whether symptomatic or asymptomatic), SARS-CoV-2-related testing and results, and vaccination details, daily. Upon registration, app users provide demographic and clinical data including age, height, weight, sex, comorbidities (i.e., cancer, diabetes, eczema, heart disease, lung disease, kidney disease and hay fever), and healthcare worker status. The app contents can also be modified to address arising research questions. We used data from 413,148 non-healthcare worker users who answered a mental health survey between February and April 2021, and reported a SARS-CoV-2 test result. Healthcare workers were excluded from this analysis due to their likely differing pandemic experience.

### Measures

Between 23 February 2021 and 12 April 2021 app contributors were invited to answer a survey about their mental health. Anxiety/depression symptoms were measured using the Generalised Anxiety Disorder assessment-2 (GAD-2)^18^ and the Patient Health Questionnaire-2 (PHQ-2)^19^. These measures examine symptoms in the preceding two weeks, each using two questions. For each question regarding frequency of a proposed situation/feeling, users can answer “Not at all”, “Several days”, “More than half the days” or “Nearly every day”.

Each answer scores from 0 for “Not at all” to 3 for “Nearly every day”. Each questionnaire has a score ranging from 0-6. Previous studies have shown an optimal cut-off point for possible anxiety or depression disorder of ≥3, yielding a sensitivity of >80%^18,19^. A binary outcome variable was created by grouping those who scored ≥3 in either GAD-2 or PHQ-2, or <3.

Logistic regression was used to study whether mental health status was associated with a positive SARS-CoV-2 test result. We adjusted for age, sex, body mass index (BMI) groups (underweight, overweight, obese, with normal BMI (18.5-24.9) as the reference), and comorbidities including learning disabilities; and applied inverse probability weighting for the probability of getting tested for SARS-CoV-2. Data are presented using descriptive statistics. Additionally, logistic regressions were employed stratifying by age groups (18-30, 31-40, 41-50, 51-60, 61-70, >70). A sensitivity analysis was performed stratifying by presence of pre-pandemic mental health disorders (see Supplementary Materials).

Further analyses in SARS-CoV-2 affected individuals assessed for association with days since infection confirmation and anxiety/depression symptoms, using days between positive test date and date of survey, grouped into: <30, 30-60, 60-90, 90-120, and >120 days.

Individuals reporting a positive test result >120 days before answering the survey served as reference.

Data were extracted and pre-processed with ExeTera^20^, a Pandas-like library developed at KCL, and statistical analysis was performed using Python (Pandas, NumPy and SciPy).

Ethical approval for use of the app for research purposes in the UK was granted by the KCL Ethics Committee (review reference LRS-19/20-18210); all users provided consent for non-commercial use.

## Results

Between 23 February and 12 April 2021, 421,977 non-healthcare workers (aged 18-99 years, BMI 15-55) answered the mental health survey and logged a SARS-CoV-2 test result (386,150 negative, 35,827 positive). 26,998 positive tests were PCR or lateral flow results; positive antibody tests (8,829) were excluded as time of infection was unknown.

26.4% (109,116) of participants scored ≥3 in GAD2 and/or PHQ2. Participants with anxiety/depression symptoms were younger, had more comorbidities, and were more often female, compared to unaffected individuals. Among those predicted to have anxiety or depression (based on a score of ≥3 on GAD-2 or PHQ-2), 38.06% (41,525) reported a previous diagnosis of a pre-pandemic mental health disorder and 5.79% (6,320) a learning disability. The study population’s demographic characteristics are presented in **Table 1**.

**Table 1:** Demographic characteristics table

|  | Anxiety or Depression symptoms<br>YES<br>n = 109116 (26.4%) | Anxiety or Depression symptoms<br>NO<br>n = 304032 (73.6%) | Total<br>n = 413148 |
| --- | --- | --- | --- |
| Age, years (mean, standard deviation) | 50.7 (13.9) | 55.6 (12.9) | 54.1 (13.4) |
| Underweight (BMI < 18.5) | 1459 (1.34%) | 3190 (1.05%) | 4649 (1.13%) |
| Normal weight (BMI 18.6 - 24.9) | 40816 (37.41%) | 134695 (44.30%) | 175511 (42.48%) |
| Overweight (BMI 25 - 29.9) | 36012 (33.00%) | 106455 (35.01%) | 142467 (34.48%) |
| Obese (BMI > 30) | 30829 (28.25%) | 59692 (19.63%) | 90521 (21.91%) |
| Has Comorbidity | 22006 (20.17%) | 52558 (17.29%) | 74564 (18.05%) |
| Has No Comorbidity | 87110 (79.83%) | 251474 (82.71%) | 338584 (81.95%) |
| Previously diagnosed with mental health condition | 41525 (38.06%) | 57067 (18.77%) | 98592 (23.86%) |
| No previous mental health condition | 67591 (61.94%) | 246965 (81.23%) | 314556 (76.14%) |
| Learning disability YES | 6320 (5.79%) | 9987 (3.28%) | 16307 (3.95%) |
| Learning disability NO | 102796 (94.21%) | 294045 (96.72%) | 396841 (96.05%) |
| Female | 80446 (73.73%) | 200563 (65.97%) | 281009 (68.02%) |
| Male | 28670 (26.27%) | 103469 (34.03%) | 132139 (31.98%) |
| Negative COVID-19 Test | 100897 (92.47%) | 285253 (93.82%) | 386150 (93.47%) |
| Positive COVID-19 Test | 8219 (7.53%) | 18779 (6.18%) | 26998 (6.53%) |
| Positive COVID-19<br><30 days before survey | 1314 (15.99%) | 2480 (13.21%) | 3794 (14.05%) |
| Positive COVID-19<br>30-60 days before survey | 2149 (26.15%) | 5043 (26.85%) | 7192 (26.64%) |
| Positive COVID-19<br>60-90 days before survey | 1843 (22.42%) | 4505 (23.99%) | 6348 (23.51%) |
| Positive COVID-19<br>90-120 before survey | 1162 (14.14%) | 2791 (14.86%) | 3953 (14.64%) |
| Positive COVID-19<br><120 before survey | 1751 (21.30%) | 3960 (21.09%) | 5711 (21.15%) |

SARS-CoV-2 infection was associated with anxiety/depression symptoms (OR 1.08, 95% CI [1.07 –1.10], p<0.001). However, stronger associations with anxiety/depression symptoms were observed for unhealthy BMI categories (i.e., underweight, overweight, obese) with odds ratios of 1.26 ([1.22 –1.30], p<0.001), 1.21 ([1.20 –1.22], p<0.001), and 1.61 ([1.59 – 1.62], p<0.001), respectively. Participants reporting one or more comorbidities (OR 1.25, 95% CI [1.24,1.26], p<0.001), and those with learning disabilities (OR 1.35, 95% CI [1.33 – 1.37], p<0.001) were more likely to have anxiety/depression symptoms. Individuals reporting a previously diagnosed mental health condition had the highest odds of reporting anxiety/depression symptoms (OR 2.26, 95% CI [2.24 – 2.28], p<0.001) (**Figure 1)**.

**Figure 1:**
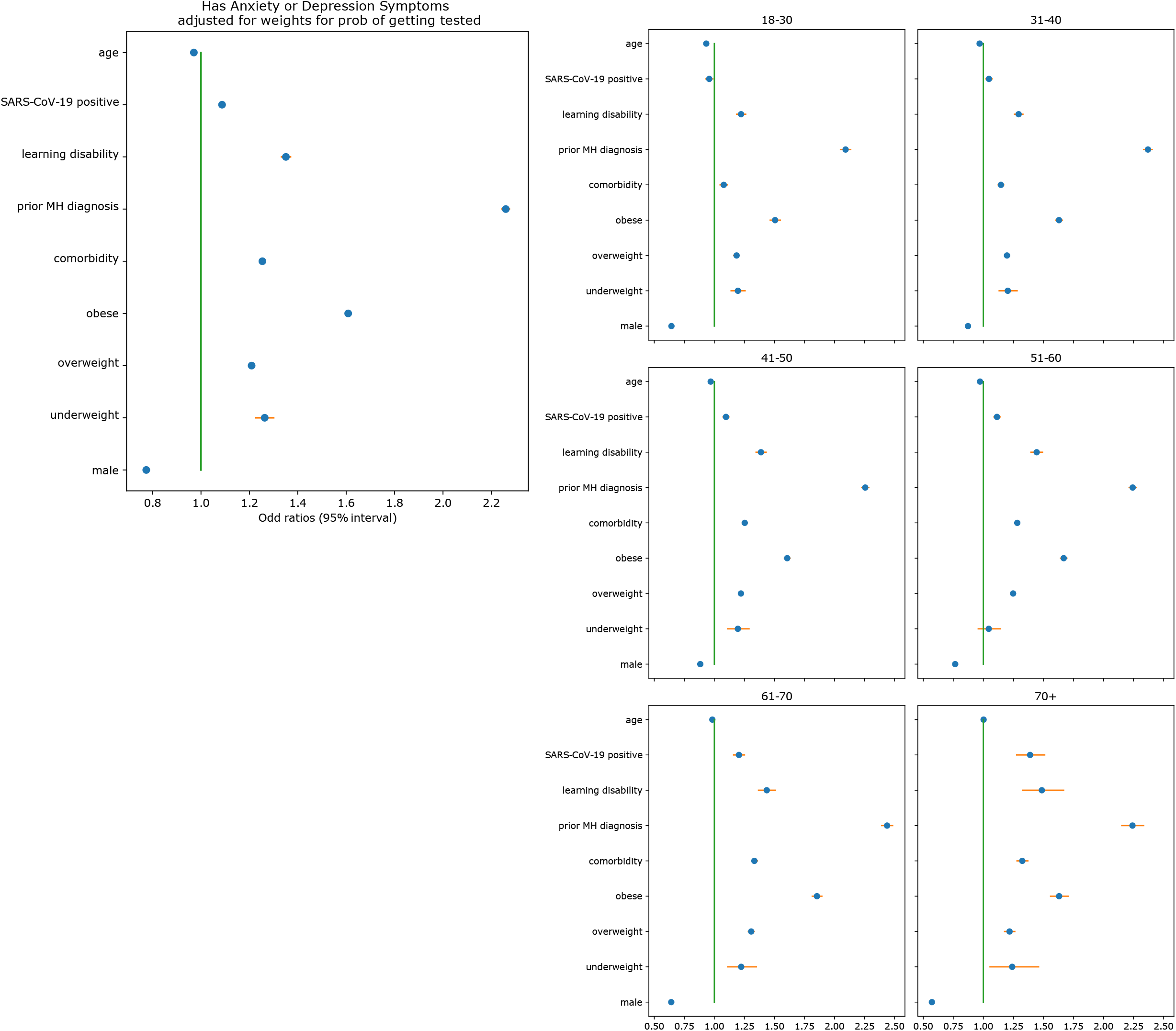
Association between age, BMI, male sex, comorbidities, a previous diagnosis of a mental health condition, learning disabilities, a positive SARS-CoV-2 test result (PCR and lateral flow), and the odds ratio of anxiety/depression symptoms suggested by the results of the mental health survey.

We observed no significant difference in the small overall increased odds of anxiety/depression symptoms with SARS-CoV-2 infection in those with a history of prior mental health conditions (OR 1.09, 95% CI [1.06 – 1.12], p <0.001); and those without such prior history of 1.09, 95% CI [1.07 – 1.10], p<0.001).

Stratification by age group showed no association between a positive SARS-CoV-2 test and anxiety/depression symptoms in young groups (<40 years). Other variables (sex, comorbidities, BMI) were consistent in age-stratified analyses. Finally, in the 26,998 cases positive for SARS-CoV-2 by PCR and lateral flow, we tested whether elapsed time after a positive test affected mental health. The relationship between SARS-CoV-2 and depression/anxiety symptoms changed over time, with increased risk of anxiety/depression symptoms in those diagnosed <30 days compared to those diagnosed >120 days prior to the survey (OR 1.15, 95% CI [1.10 -1.2], p<0.001).

## Discussion

In this large, community-based study, we report a small positive association between SARS-CoV-2 infection and anxiety/depression symptoms. However, this was dwarfed by associations with the known risk factors BMI, sex, and comorbidities. Results were robust to sensitivity analyses stratifying by prior mental health disorder diagnoses. Further, no association between SARS-CoV-2 infection and anxiety/depression symptoms was found in younger age groups (<40 years).

Association between SARS-CoV-2 infection and anxiety/depression symptoms changed over time, with the strongest association in those infected <30 days prior to the survey, suggesting a short-term effect of infection on mental health only. It is possible that other factors affecting mental health which were changing over the pandemic (e.g., lockdown) may moderate an effect of elapsed time since SARS-CoV-2 infection on mental health^21^.

Overall prevalence of anxiety/depression symptoms in our study (26.4%) is slightly increased compared to pre-pandemic levels of mental health issues in the UK general population assessed by the UK Household Longitudinal Study with the GHQ-12 questionnaire (18.9% in 2018^22^) but broadly comparable to the level seen in April 2021 (27.3%^22^). This previous 2021 study did not explore any relationship with SARS-CoV-2 infection. A recent analysis of 1112 subjects experiencing probable COVID-19 symptoms suggested a positive association between COVID-19 and anxiety/depression symptoms 1-7 months after suggested infection (OR 1.31 -1.47)^23^. Our study benefits from a much larger sample size of tested participants.

Our study has several limitations. Data are self-reported using a mobile app, and may disproportionately represent more affluent populations. We only had one time point of mental health data collection, limiting our ability to test if associations changed as the pandemic progressed. Additionally, although we applied weighting for the probability of being tested for the virus, results referring to time since testing might be still biased due to limited testing capacity early in the pandemic. As in any study assessing mental health through questionnaires, selection bias (whereby mental health influences who responds) and reporting bias (relating to perception, and/or influence of a ‘valid’ reason to report) may limit the validity of our results. Further analyses of longitudinal datasets with different reporting structures are warranted.

## Conclusion

This study suggests a weak association between SARS-CoV-2 infection and anxiety/depression symptoms, especially in adults >40 years, which is small relative to known risk factors such as previous medical or mental health conditions and/or unhealthy BMI. The association was most evident in recently infected individuals. This suggests that an effect of SARS-CoV-2 on mental health may be only of short duration. Further exploration may help to understand factors that will improve mental health after SARS-CoV-2 infection.

## Data Availability

Anonymised research data are shared with third parties via the centre for Health Data Research UK (HDRUK.ac.uk). US investigators are encouraged to coordinate data requests through the COPE Consortium (www.monganinstitute.org/cope-consortium). Data updates can be found on https://covid.joinzoe.com

https://www.hdruk.ac.uk/

https://www.monganinstitute.org/cope-consortium

https://covid.joinzoe.com

## Acknowledgements

This work is supported by the Wellcome Engineering and Physical Sciences Research Council (EPSRC) Centre for Medical Engineering at King’s College London (WT 203148/Z/16/Z) and the UK Department of Health via the National Institute for Health Research (NIHR) comprehensive Biomedical Research Centre (BRC) award to Guy’s & St Thomas’ NHS Foundation Trust in partnership with King’s College London and King’s College Hospital NHS Foundation Trust, the Medical Research Council (MRC) and British Heart Foundation. SO and MM are supported by the UK Research and Innovation London Medical Imaging & Artificial Intelligence Centre for Value Based Healthcare, and the Wellcome Flagship Programme (WT213038/Z/18/Z). This research was funded in part by the Wellcome Trust [WT213038/Z/18/Z] and [WT203148/Z/16/Z]. For the purpose of open access, the author has applied a CC BY public copyright license to any Author Accepted Manuscript version arising from this submission. EM is funded by an MRC Skills Development Fellowship Scheme at KCL. CHS, EJT and CJS are supported by the National Core Studies, an initiative funded by UKRI, NIHR and the Health and Safety Executive, and funded by the Medical Research Council (MC_PC_20030). CHS is also supported by an Alzheimer’s Society Junior Fellowship (AS-JF-17-011). Zoe Limited supported all aspects of building and running the app and service to all users worldwide. ATC is the Stuart and Suzanne Steele MGH Research Scholar. LHN is supported by an NIH K23DK125838 award, the American Gastroenterological Association Research Scholars Award, and the Crohn’s and Colitis Foundation Career Development Award. DAD and LHN are supported by the AGA-Takeda COVID-19 Rapid Response Research Award (AGA2021-5102). DAD is supported by NIH/NIDDK K01DK120742. ATC, LHN, and DAD are supported by the Massachusetts Consortium on Pathogen Readiness (MassCPR). We express our sincere thanks to all the participant users of the app, including study volunteers enrolled in cohorts within the Coronavirus Pandemic Epidemiology (COPE) consortium. We thank the staff of Zoe Limited, the Department of Twin Research at King’s College London, the Clinical & Translational Epidemiology Unit at Massachusetts General Hospital, Researchers and staff at Lund University in Sweden for their tireless work in contributing to the running of the study and data collection.

