## Supplemental Material for "Anxiety and depression symptoms after COVID-19 infection: results from the COVID Symptom Study app"

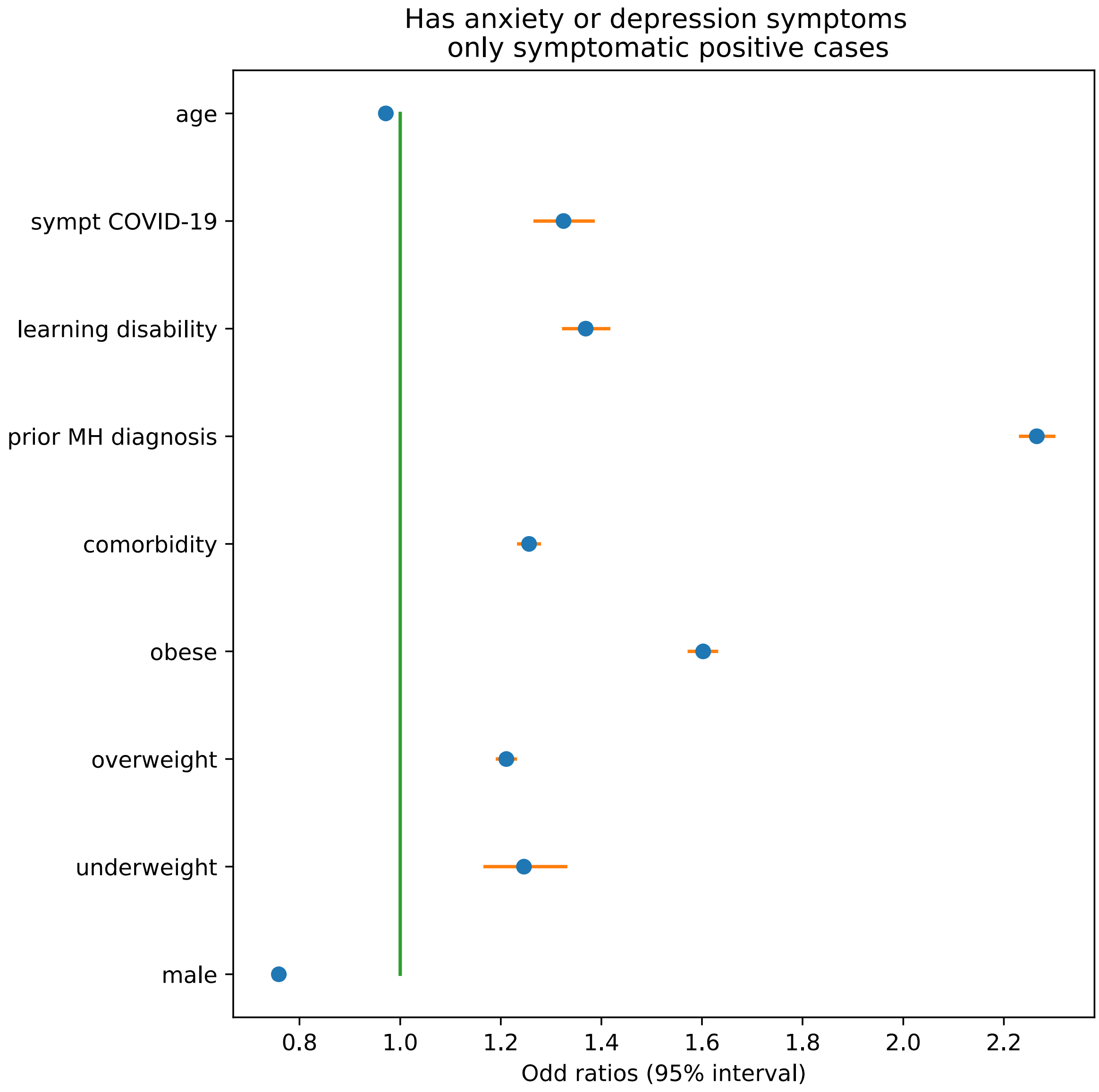


**Supplementary Figure 1**: Association between age, BMI, male sex, comorbidities, a previous diagnosis of a mental health (MH) condition, learning disabilities, a positive SARS-CoV-2 test result (PCR and lateral flow), and the odds ratio of anxiety/depression symptoms suggested by the results of the mental health survey. Only symptomatic cases were considered in this sensitivity analysis.

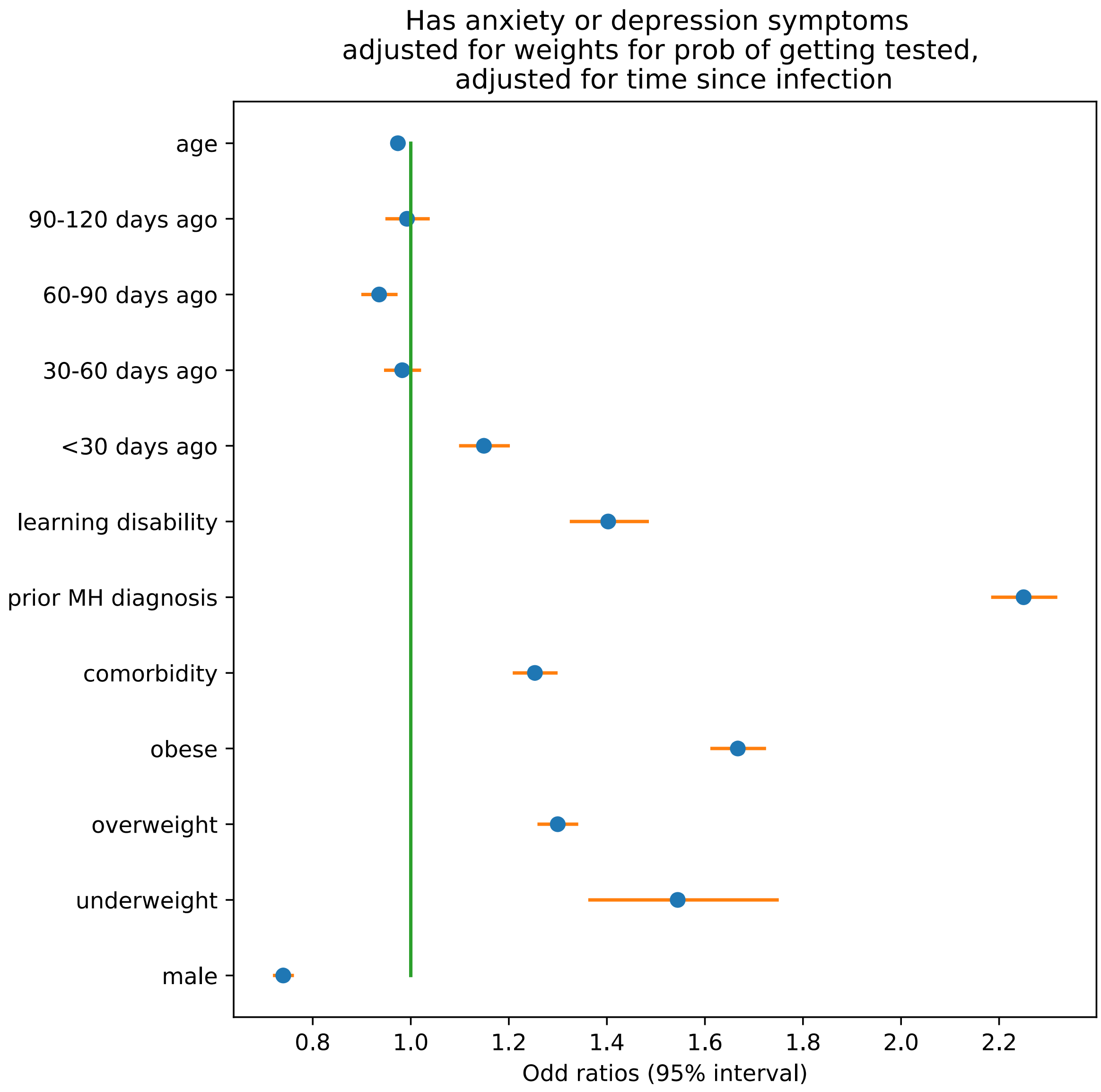


**Supplementary Figure 2**: Association between age, BMI, male sex, comorbidities, a previous diagnosis of a mental health (MH) condition, learning disabilities, time since infection occured, and the odds ratio of anxiety/depression symptoms suggested by the results of the mental health survey. This sensitivity analysis was performed on users with a positive SARS-CoV-2 test result (PCR and lateral flow) only.

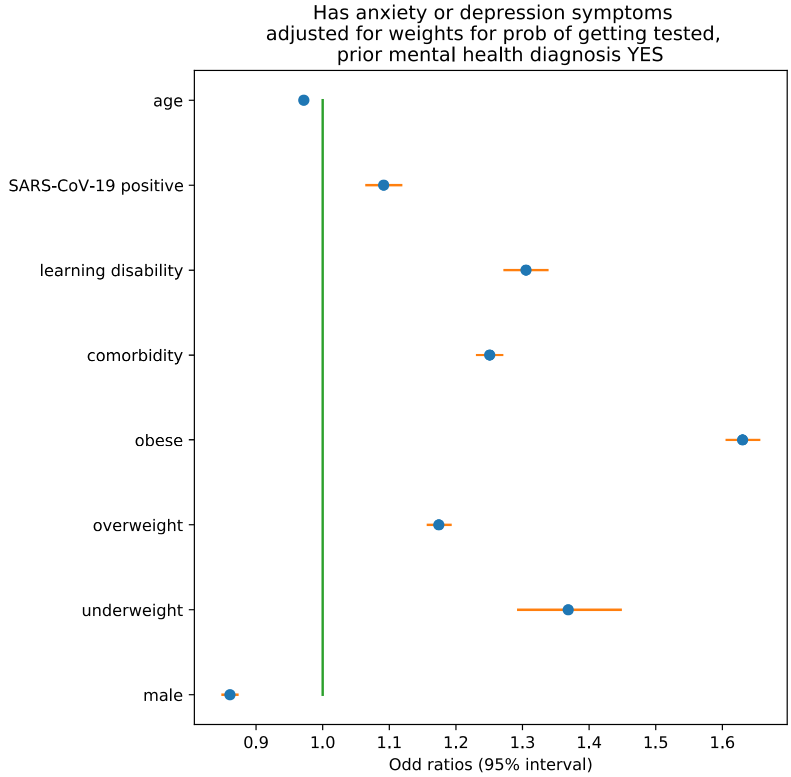


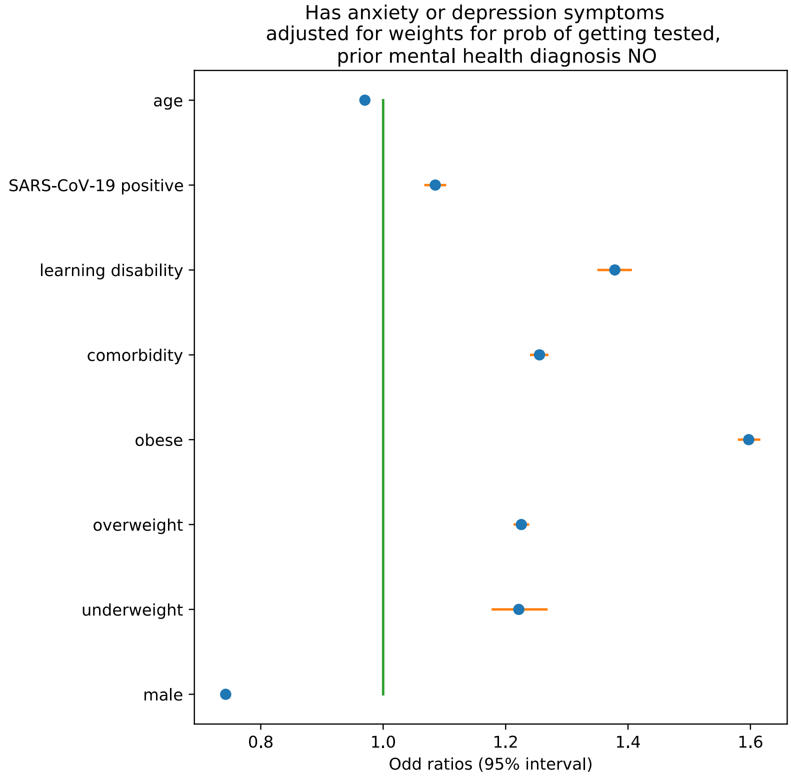


**Supplementary Figure 3**: Association between age, BMI, male sex, comorbidities, learning disabilities, a positive SARS-CoV-2 test result (PCR and lateral flow), and the odds ratio of anxiety/depression symptoms suggested by the results of the mental health survey stratified by prior mental health condition.

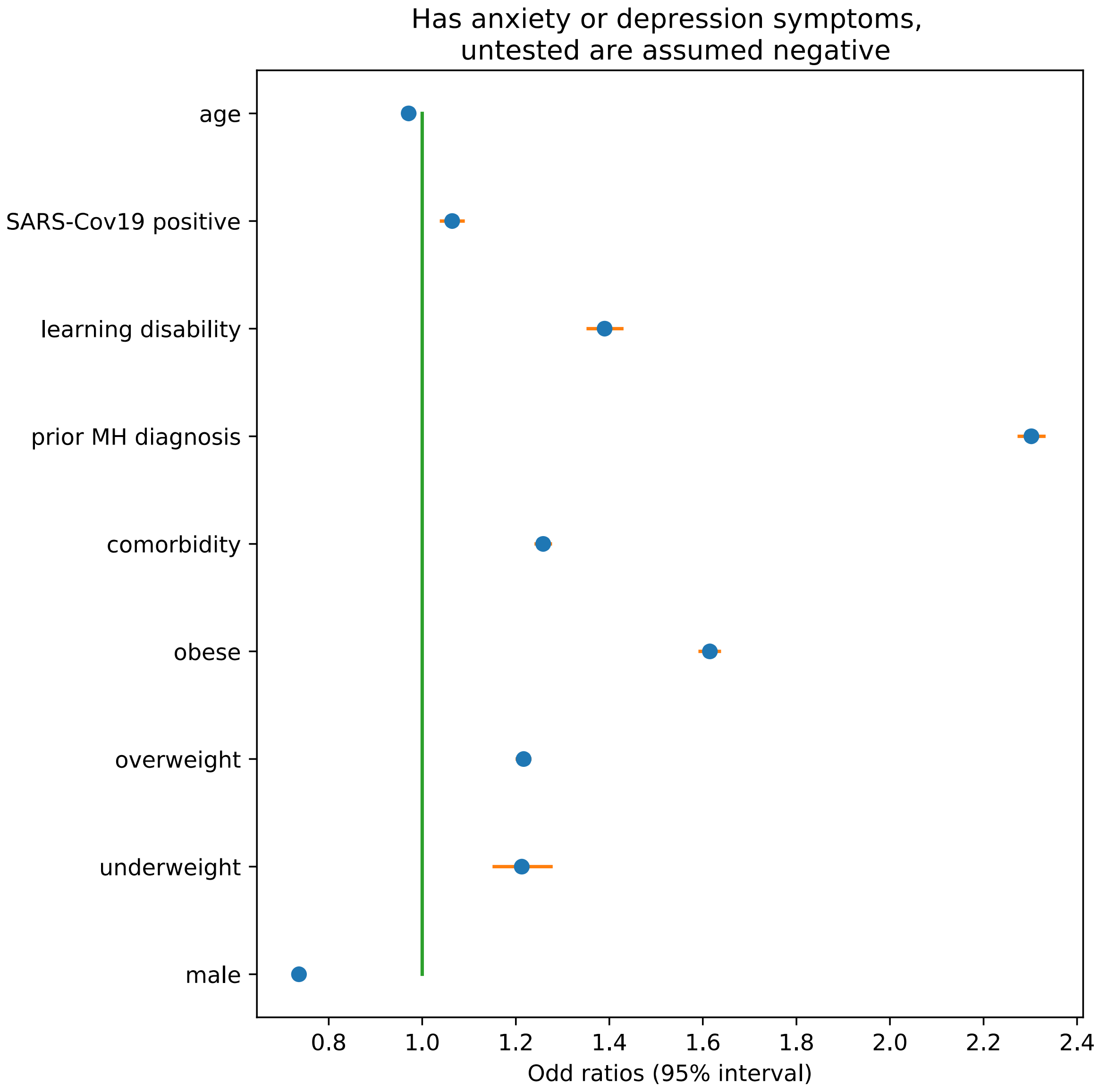


**Supplementary Figure 4**: Association between age, BMI, male sex, comorbidities, a previous diagnosis of a mental health (MH) condition, learning disabilities, a positive SARS-CoV-2 test result (PCR and lateral flow), and the odds ratio of anxiety/depression symptoms suggested by the results of the mental health survey. Untested cases were assumed negative test results in this sensitivity analysis.
